# Care Home Residence and Carbapenemase-Producing Enterobacterales Positivity: A Matched Case-Control Study

**DOI:** 10.64898/2026.07.02.26357111

**Authors:** Harmony Owhotake, Jossy Ashlin, Stefano Oggiano, Aiden J. Plant

## Abstract

**Background:** Carbapenemase-producing Enterobacterales (CPE) remain a major infection prevention and control challenge. Although care home residence is frequently perceived as a risk factor for CPE carriage, its independent association with CPE positivity remains uncertain.

**Objective:** To investigate the relationship between care home residence on admission and CPE positivity among patients undergoing CPE screening.

**Methods:** A retrospective matched case-control study was conducted at a single NHS acute hospital in England. Adult patients with laboratory-confirmed CPE positivity from screening samples between 1 November 2022 and 1 November 2025 were matched to CPE-negative controls at a ratio of up to 1:4 based on ward, specimen year and age within ±5 years. Conditional logistic regression was used to assess the association between care home residence and CPE positivity. An adjusted model included previous hospital admission within 12 months.

**Results:** A total of 108 CPE-positive cases were successfully matched to 412 controls. Care home residence was identified in 14 (13.0%) cases and 49 (11.9%) controls. In the matched conditional logistic regression model, care home residence was not associated with CPE positivity (OR 1.15, 95% CI 0.58–2.28; p=0.690) and remained non-significant after adjustment (aOR 1.32, 95% CI 0.66–2.64; p=0.439).

**Discussion:** Care home residence was not independently associated with CPE positivity in this low-prevalence setting.

**Significance and impact:** The findings do not support the use of care home residence alone to guide CPE screening. Further multicentre studies are required to clarify the contribution of care home residence to CPE epidemiology.

## Introduction

Carbapenemase-producing Enterobacterales (CPE) are recognized as a critical threat to patient safety and public health due to their resistance to last-line antibiotics, limited therapeutic options with increased mortality, and high potential for healthcare-associated transmission (Paterson and Doi, 2015; UK Health Security Agency, 2022). The global spread of carbapenemase genes has been identified as a priority area for antimicrobial resistance (AMR) containment, with healthcare settings playing a central role in amplification and dissemination (World Health Organization, 2024)

In the United Kingdom, CPE pose a particular challenge for infection prevention and control (IPC) teams. Colonised patients may be asymptomatic, yet act as reservoirs for onward transmission within hospitals, particularly in high-risk clinical areas (Gagliotti *et al*., 2013; Marimuthu *et al*., 2022; Kim *et al*., 2023). As a result, timely identification of patients at increased risk of CPE carriage on admission is essential to inform screening, isolation, and cohorting strategies (UK Health Security Agency, 2022)

National screening guidance focuses on epidemiological risk factors such as recent overseas healthcare exposure, known previous CPE carriage, or contact with confirmed cases or outbreaks (UK Health Security Agency, 2022). While these criteria are evidence-based, they may not capture all patients at elevated risk within local healthcare systems. Care home residents frequently have complex healthcare needs, recurrent hospital admissions, frequent antibiotic exposure, and close contact with healthcare services, all of which may increase the likelihood of colonization with resistant organisms (Chen *et al*., 2021).

Care homes are recognised reservoirs of antimicrobial-resistant organisms, with evidence showing associations with Extended-spectrum β-lactamase (ESBL)-producing Enterobacterales (Ben-Ami *et al*., 2009). Whether a similar relationship exists for CPE remains uncertain. In a previous evaluation of our Trust’s CPE screening programme, the highest screening yield was observed among patients admitted from a local care home with a known CPE outbreak (Owhotake, Oggiano and Plant, 2026). This finding raised the question of whether care home residence itself was independently associated with CPE positivity, or whether the observed risk reflected local epidemiological clustering. This study, therefore, aimed to determine whether care home residence was independently associated with CPE positivity.

## Methods

### Study design and setting

A retrospective matched case-control study was conducted within a single NHS acute hospital in England. The hospital was a 550-bed district general hospital in the West Midlands region, providing acute secondary care services to approximately 294,800 residents (*Walsall Demographics* | *Walsall Council*, no date). The setting had a low CPE prevalence with a detection rate of 0.6 per 1,000 admissions and a screening positivity of 2.1% (Owhotake, Oggiano and Plant, 2026).

The study evaluated patients who underwent CPE screening as part of the Trust’s routine infection prevention and control surveillance programme from November 1^st^ 2022 to November 1^st^ 2025. Patients meeting the Trust’s risk-based CPE screening criteria underwent screening in accordance with local policy. Screening eligibility included recognised risk factors for CPE carriage, such as previous healthcare exposure and known CPE risk factors, and has been described in detail elsewhere (Owhotake, Oggiano and Plant, 2026).

### Study population

Cases were defined as adult patients (≥18 years) with laboratory-confirmed CPE positivity identified from screening specimens only during the study period (1 November 2022 and 1 November 2025). Patients with a documented history of CPE positivity within 12 months preceding admission were excluded, in accordance with the Trust’s policy whereby patients are considered to have a recent history of CPE carriage for 12 months following a positive result. Controls were adult patients who underwent CPE screening during the same period with a negative result.

Screening records were consolidated to patient-level entries. Patients with CPE positivity were counted once at the time of their first positive result. For CPE-negative patients, multiple screening episodes and admissions during the study period were consolidated into a single patient-level record using the earliest negative screening result. Patients included as controls had no documented CPE-positive screening result during the study period. Individuals initially identified through a negative screening result who subsequently became CPE positive were reclassified as cases and excluded from the control dataset prior to matching.

### Screening programme

During the study period, CPE screening was undertaken according to local policy and included patients meeting predefined epidemiological risk criteria (Owhotake, Oggiano and Plant, 2026). Screening occurred on admission or during admission when one or more screening criteria were identified. Screening specimens comprised rectal swabs or faecal samples, depending on clinical opportunity. A case of CPE was defined as the isolation of an Enterobacterales species harbouring a carbapenemase enzyme.

Laboratory processing was performed using standard culture techniques. Organism identification was undertaken using matrix-assisted laser desorption/ionisation time-of-flight (MALDI-TOF) mass spectrometry, while carbapenemase genes were confirmed by multiplex polymerase chain reaction (PCR) targeting Verona integron-encoded metallo-β-lactamase (VIM), imipenemase metallo-β-lactamase (IMP), New Delhi metallo-β-lactamase (NDM), Klebsiella pneumoniae carbapenemase (KPC), and oxacillinase-48 (OXA-48).

### Matching strategy

Controls were matched to cases at a ratio of up to 1:4 based on ward, specimen year, and age (±5 years). Ward referred to the patient’s location at the time of specimen collection and included both general inpatient wards and critical care areas where applicable. A 1:4 matching ratio was selected to maximise statistical efficiency while retaining all eligible cases. Matching was performed without replacement. Cases for whom no eligible controls could be identified were excluded from matched analyses. This approach was chosen to minimise confounding related to the clinical setting and age-related risk.

### Variables

The primary exposure was care home residence (yes/no), defined as residence in a long-term care facility immediately prior to hospital admission. The primary outcome was CPE positivity. Additional variables through manual record review included previous hospital admission within 12 months, documented CPE contact exposure and care home name to assess clustering, if required.

### Statistical analysis

Descriptive statistics were used to summarise baseline characteristics. Conditional logistic regression was employed to estimate the association between care home residence and CPE positivity while accounting for the matched study design. An adjusted conditional logistic regression model was subsequently fitted, including previous hospital admission within 12 months. Odds ratios (ORs) and adjusted odds ratios (aORs) are presented with 95% confidence intervals (CIs). Statistical significance was defined as p<0.05. Analyses were performed using Python (version 3.13.5).

### Ethical considerations

This study used fully anonymised, routinely collected surveillance data and met criteria for service evaluation.

## Results

### Study population

A total of 140 CPE-positive patients was identified during the study period. Following exclusion of 5 paediatric patients, 1 patient with known CPE positivity from another hospital within 12 months prior to admission, 20 patients who could not be matched to eligible controls and 6 clinical specimens, 108 cases were successfully matched to 412 controls and included in the final analysis. Patient characteristics are shown in Table I.

**Table I.**
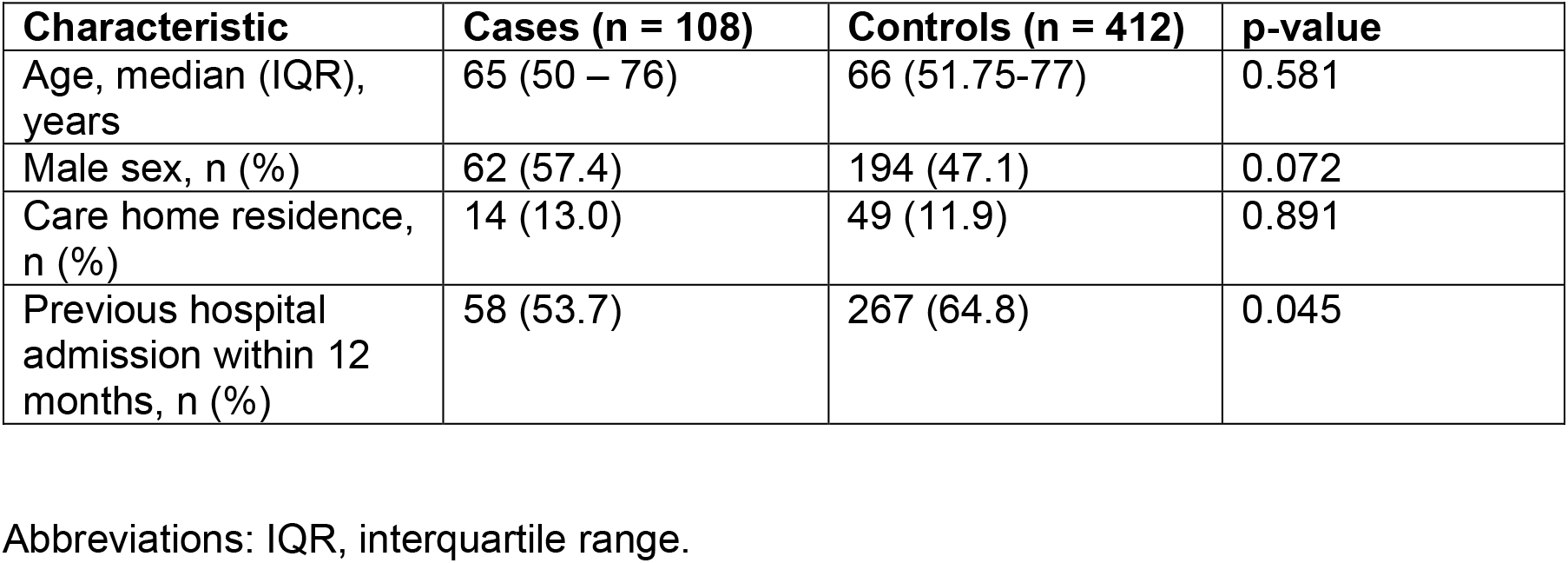
Baseline characteristics.

### Primary analysis

In the matched conditional logistic regression model, care home residence was not associated with CPE positivity (OR 1.15 95% CI 0.58 – 2.28; p-value = 0.690).

### Adjusted analysis

After adjustment for previous hospital admission within 12 months, care home residence remained unassociated with CPE positivity (aOR 1.32 95% CI 0.66-2.64; p-value = 0.439). Full model results are presented in Table II.

**Table II.** Adjusted conditional logistic regression model for CPE positivity.

| **Variable** | **aOR** | **95% CI** | **p-value** |
| --- | --- | --- | --- |
| Care home residence | 1.32 | 0.66-2.64 | 0.439 |
| Previous hospital admission within 12 months | 0.61 | 0.39-0.96 | 0.033 |

Abbreviations: aOR, adjusted odds ratio; CI, confidence interval; OR, odds ratio

### Care home clustering

Fourteen CPE-positive care home residents originated from ten separate care homes.

One care home contributed five cases, while the remaining nine care homes contributed a single case each. Due to the small number of care-home-associated cases, no formal assessment of facility-level clustering was undertaken.

## Discussion

In this matched case-control study, care home residence was not independently associated with CPE positivity in both the unadjusted and adjusted analyses. Although care home residents represented a notable proportion of CPE-positive patients, a similar prevalence was observed among matched controls. This suggests that care home residence alone may have limited value as an independent risk marker for CPE positivity in this low-prevalence setting.

Previous studies have identified care homes as reservoirs of antimicrobial-resistant organisms, including ESBL-producing Enterobacterales (Ben-Ami *et al*., 2009). In addition, our previous evaluation of the Trust’s CPE screening programme found the highest screening yield among patients admitted from a local care home with a known CPE outbreak (Owhotake, Oggiano and Plant, 2026). Taken together, these findings suggest an important distinction between care home residence in general and exposure to a care home with known or suspected CPE activity. The latter may remain relevant to local IPC risk assessment, even where care home residence alone is not independently associated with CPE positivity.

One care home contributed five of the fourteen CPE-positive care home residents identified in the study. However, the small number of care-home-associated cases prevented formal assessment of whether CPE risk differed between individual care homes.

This study has several limitations. It was conducted within a single hospital, limiting generalisability. As a retrospective study, data collection was limited to information routinely documented within clinical and IPC records. Detailed clinical data on comorbidity, invasive devices and antibiotic exposure were unavailable, and residual confounding related to illness severity cannot be excluded. Although all eligible cases identified during the study period were included, the relatively small number of care-home-associated cases may have limited the ability to detect small differences between groups. Information on documented CPE contact exposure was available; however, the prevalence of documented contact exposure was similar among cases and controls and was therefore not included in the final adjusted analysis. Controls were represented by their earliest negative screening episode, whereas cases were represented by their first positive screening episode. Because some cases may have undergone one or more negative screening episodes before their first positive result, the screening episode used for analysis may not have represented comparable time points for cases and controls.

Consequently, variables related to screening timing were not included in the primary interpretation of the findings. Fourteen care-home-associated cases were identified across ten care homes, limiting the ability to formally assess facility-level clustering. In addition, the study was conducted in a low-prevalence setting, and the findings may not be generalisable to settings with higher endemic CPE prevalence.

These findings do not support the use of care home residence alone as a criterion for CPE screening. However, local epidemiological intelligence, including identification of care home-associated clusters or known CPE activity within specific facilities, may remain important when assessing the need for targeted IPC interventions. Further multicentre studies are warranted to explore the generalisability of these findings and their implications for screening policy.

## Conclusion

Care home residence was not independently associated with CPE positivity in this matched case-control study before and after adjustment for previous hospital admission. Although care homes are recognised reservoirs of antimicrobial-resistant organisms, residence in a care home alone did not identify patients at increased risk of CPE positivity in this low-prevalence setting. Screening decisions should therefore be informed by established risk factors and local epidemiological intelligence, including known or suspected CPE activity within specific facilities.

## Data Availability

The data is not publicly available due to organisational governance restrictions, but may be available from the corresponding author on reasonable request.

## Notes

**Declaration of conflicting interest:** The authors declare no conflicts of interest.

### Competing Interest Statement

The authors have declared no competing interest.

### Author Declarations

The study was conducted within Walsall Healthcare NHS Trust, England, using fully anonymised routinely collected surveillance data. The NHS Health Research Authority decision tool determined that NHS Research Ethics Committee review was not required for this study.

